# Non-alcoholic fatty liver disease (NAFLD) and risk of hospitalization for Covid-19

**DOI:** 10.1101/2020.09.01.20185850

**Authors:** Carolyn T. Bramante, Christopher J. Tignanelli, Nirjhar Dutta, Emma Jones, Leonardo Tamaritz, Jeanne Clark, Genevieve Melton-Meaux, Michael Usher, Sayeed Ikramuddin

## Abstract

**Background:** Covid-19 disease causes significant morbidity and mortality through increase inflammation and thrombosis. Non-alcoholic fatty liver disease and non-alcoholic steatohepatitis are states of chronic inflammation and indicate advanced metabolic disease. We sought to understand the risk of hospitalization for Covid-19 associated with NAFLD/NASH.

**Methods:** Retrospective analysis of electronic medical record data of 6,700 adults with a positive SARS-CoV-2 PCR from March 1, 2020 to Aug 25, 2020. Logistic regression and competing risk were used to assess odds of being hospitalized. Additional adjustment was added to assess risk of hospitalization among patients with a prescription for metformin use within the 3 months prior to the SARS-CoV-2 PCR result, history of home glucagon-like-peptide 1 receptor agonist (GLP-1 RA) use, and history of metabolic and bariatric surgery (MBS). Interactions were assessed by gender and race.

**Results:** A history of NAFLD/NASH was associated with increased odds of admission for Covid-19: logistic regression OR 2.04 (1.55, 2.96, p<0.01), competing risks OR 1.43 (1.09-1.88, p<0.01); and each additional year of having NAFLD/NASH was associated with a significant increased risk of being hospitalized for Covid-19, OR 1.86 (1.43-2.42, p<0.01). After controlling for NAFLD/NASH, persons with obesity had decreased odds of hospitalization for Covid-19, OR 0.41 (0.34-0.49, p<0.01). NAFLD/NASH increased risk of hospitalization in men and women, and in all racial/ethnic subgroups. Mediation treatments for metabolic syndrome were associated with non-significant reduced risk of admission: OR 0.42 (0.18-1.01, p=0.05) for home metformin use and OR 0.40 (0.14-1.17, p=0.10) for home GLP-1RA use. MBS was associated with a significant decreased risk of admission: OR 0.22 (0.05-0.98, p<0.05).

**Conclusions:** NAFLD/NASH is a significant risk factor for hospitalization for Covid-19, and appears to account for risk attributed to obesity. Treatments for metabolic disease mitigated risks from NAFLD/NASH. More research is needed to confirm risk associated with visceral adiposity, and patients should be screened for and informed of treatments for metabolic syndrome.

**Key Questions:** *Question:* Does NAFLD/NASH independently increase risk for poor outcomes from Covid-19?

*Findings:* In this observational study, a history of NAFLD/NASH was associated with a significantly increased odds of hospitalization. Metabolic surgery was protective against admission in persons with NAFLD/NASH and Covid-19. Metformin and glucagon like peptide 1 receptor agonists were associated with non-significant protecting against admission.

*Meaning:* Treatment for metabolic syndrome greatly reduce the elevated risk of hospitalization for Covid-19 among persons with NAFLD/NASH.

## Background

Hospitalizations for Covid-19 disease continue to disrupt thousands of lives.^1^ Obesity and metabolic disease appear to be the most significant modifiable risk factors for poor outcomes from Covid-19.^2,3^ Hepatic steatosis (non-alcoholic fatty liver disease, NAFLD, and non-alcohol steatohepatitis [NASH]) is evidence of visceral adiposity, advanced metabolic disease, and overt inflammation.^4^ Given Covid-19’s pathophysiology through inflammation, NAFLD/NASH may put patients at even higher risk of poor outcomes from Covid-19.^5^ Few papers have fully assessed risk for poor outcomes from Covid-19 associated with NAFLD/NASH, and factors that might reduce that risk.

We conducted a retrospective analysis of individuals with SARS-CoV-2 infection and their likelihood of being admitted for Covid-19 with the following objectives: Our primary objective was to quantify the risk for hospitalization for Covid-19 based on a history of NAFLD/NASH. Our secondary objective was to assess whether known treatments for metabolic disease modified risk associated with NAFLD/NASH.^6^ Third, we were interested in whether NAFLD/NASH was associated with severe Covid-19 (intubation, mortality). Lastly, because visceral adiposity accumulates at a lower body mass index (BMI) level in men^7^ and is more prevalent among certain ethnic groups,^8-10^ we assessed interactions between gender and race/ethnicity and NAFLD/NASH as drivers of Covid-19 outcomes.

## Methods

### Design and data source

Retrospective analysis of electronic medical record data from 12 hospitals and 56 primary care clinics.

### Population

Over 6,700 adults with positive SARS-CoV-2 PCR from March 1, 2020 to Aug 25, 2020. Inclusion criteria: patients who opted in to research. All ages included. Exclusion criteria: patients who opted out of research.

### Independent Variable

Persons with NAFLD/NASH were defined as those with ICD codes for NAFLD or NASH or a BMI >= 30kg/m^2^ and an elevated alanine aminotransferase (ALT) on 3 separate dates. This definition was used rather than ICD coding because NASH and NAFLD are significantly underdiagnosed in the EHR.^11,12^

### Dependent Variable of Interest

An admission to the hospital for Covid-19 disease. Secondary outcomes were assessed for mortality, and admission to the intensive care unit (ICU).

### Analyses

Logistic regression and competing risk were used to assess odds of being hospitalized. Adjustments were made for known confounders selected by Lasso-logit to reduce the likelihood of over-fitting. Additional adjustment was added to assess risk of hospitalization among patients with a prescription for metformin use with in the 3 months prior to the SARS-CoV-2 PCR result, history of home glucagon-like-peptide 1 receptor agonist (GLP-1 RA) use, and history of metabolic and bariatric surgery (MBS). Interactions were assessed by gender and race, and subgroup analyses were subsequently conducted. In order to account for variables that can worsen NAFLD/NASH, sensitivity analysis was done excluding patients with a history of alcohol use disorder, home amiodarone or methotrexate use.^13^ Multiple imputation was used for variables with missingness over 10%. Logistic Goodness of fit was assessed using the Hosmer-Lemeshow test (p=1.00) and area under the curve (0.86) for the model pre-imputation. E-values were calculated. Statistical analyses were performed using Stata MP, version 16 (StataCorp, College Station, TX). The University of MN IRB approved this study.

## Results

### Characteristics of the Cohort

The median age was 46, (IQR 28 to 66), 44% female, and 373 (5.5%) persons had NAFLD/NASH. Fifty three percent identified as White; 22% as Black; 12% as Asian; 8% as Hispanic; 4% Declined; and 1% as “Other,” Table 1.

### Risks associated with NAFLD/NASH

A history of NAFLD/NASH was associated with increased odds of admission for Covid-19: logistic regression OR 2.04 (1.55, 2.96, p<0.01), competing risks OR 1.43 (1.09-1.88, p<0.01); and in the sensitivity analysis. Each additional year of having NAFLD/NASH was also associated with a significant increased risk of being hospitalized for Covid-19, OR 1.86 (1.43-2.42, p<0.01). After controlling for NAFLD/NASH, persons with obesity had decreased odds of hospitalization for Covid-19, OR 0.41 (0.34-0.49, p<0.01), Table 2, Figure 1.

**Figure 1.**
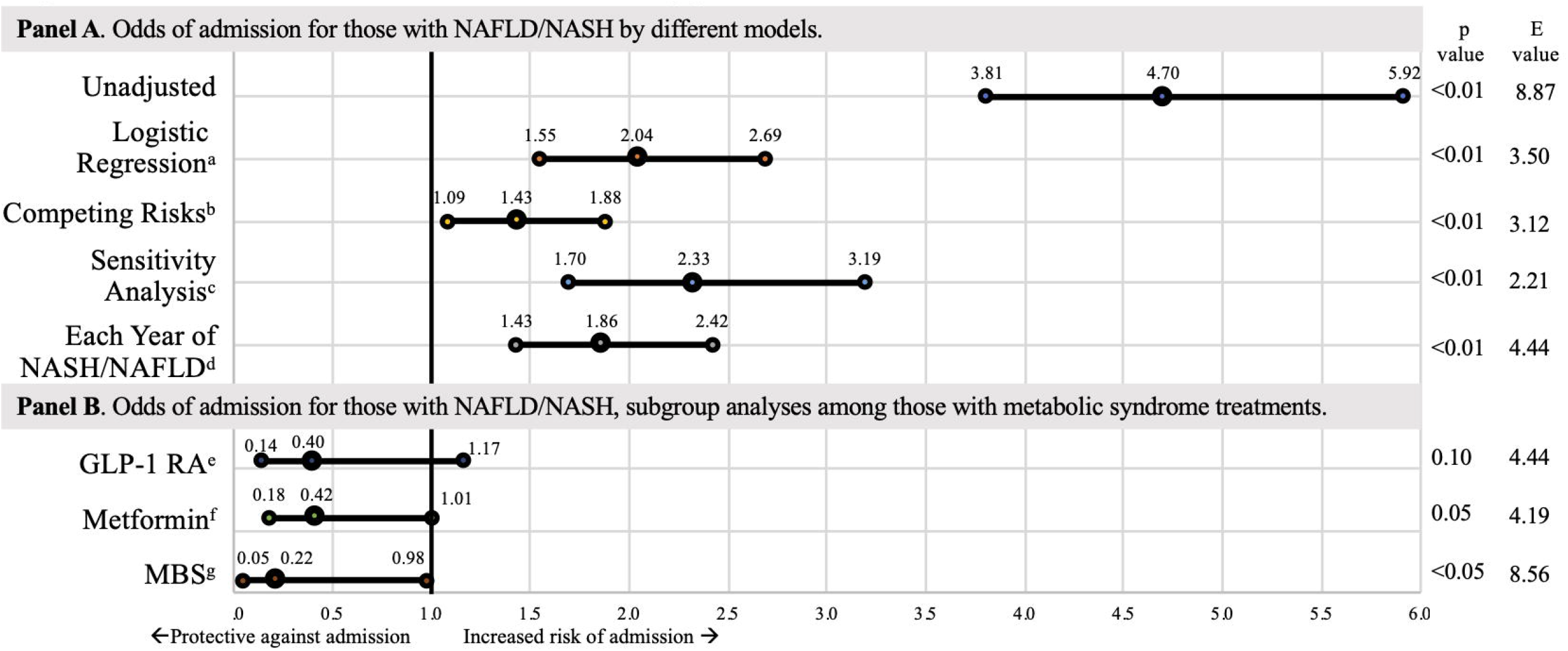
Odds of being admitted for Covid-19. The bars represent 95% confidence intervals. a Logistic regression, adjusted for age, sex, obesity, ethnicity, NAFLD/NASH, alcohol use disorder, Elixhauser comorbidity index, and home use of amiodarone, methotrexate, oral steroids, or calcium channel blockers (CCB). b Competing risk, adjusted model. c Excluding persons with alcohol use disorder, amiodarone, methotrexate, or CCB use, adjusted model. d Odds for admission for each additional year of having NAFLD/NASH. e Among those with home Glucagon-Like-Peptide-1 Receptor Agonist (GLP-1 RA), adjusted model. f Among those with home metformin use in the previous 3 months, adjusted model. g Among those with a history of metabolic and bariatric surgery (MBS), adjusted model.

### Demographic characteristics

After controlling for NAFLD/NASH, men had increased odds of admission, 1.28 (1.08-1.51, p<0.01). In subgroup analysis, NAFLD/NASH increased odds of admission in men: 2.48 (1.68-3.65, p<0.01), and among women: OR 1.71 (1.17-2.49, p=0.01). After controlling for BMI and NAFLD/NASH, persons of color still had increased risk of admission over those identifying as White. NAFLD/NASH significantly increased risk of hospitalization within each racial subgroup, with the largest increase among persons who self-identified as Black, OR 2.69 (1.40-5.16, p<0.01).

### Metabolic syndrome treatments

Persons with NAFLD/NASH who had home metformin use in the 3-months prior to SARS-CoV-2 diagnosis (n=36), had a large decrease in odds of admission with near significance, OR 0.42 (0.18-1.01, p=0.05), as did those with home GLP-1RA use, n=22, OR 0.40 (0.14-1.17, p=0.10). Persons with NAFLD/NASH who had undergone bariatric surgery, n=16, had significantly decreased odds of admission for Covid-19, OR 0.22 (0.05-0.98, p<0.05).

## Discussion

This is the first in-depth assessment of NAFLD/NASH as a risk factor for admission for Covid-19 in a large database in the US, as well as possible treatments for mitigating this risk. We found that NAFLD/NASH was a more significant risk factor for admission than age, gender, obesity, or other comorbidities. This is the first analysis to describe that after controlling for NAFLD/NASH, there was no longer an increased risk of hospitalization with obesity. This may indicate the significant role of visceral adiposity in the pathophysiology of Covid-19, which amplifies the chronic state of inflammation and hypercoagulability created by NAFLD/NASH.

NAFLD/NASH increased risk of hospitalization in both men and women. Assessing an interaction with race was done because of significant differences in outcomes in Covid-19 by race and ethnicity.^14^ NAFLD/NASH increased risk of hospitalization in all groups, but controlling for NAFLD/NASH did not eliminate differences in risk between racial/ethnic groups. Of note, persons who had self-identified as Black who had NAFLD/NASH compared to those who did not had the biggest increased odds of hospitalization. This may be due to lower amounts of visceral adiposity previously reported in Black individuals.^15^

Promisingly, we found that known treatments for metabolic syndrome and NAFLD/NASH greatly mitigated risks from Covid-19 –those with home metformin or GLP-1RA use had a non-significantly reduced odds of hospitalization, and those who had undergone bariatric surgery had a significant decrease in odds of hospitalization. CCB’s were associated with a non-significant benefit in this analysis. Findings of protective effects from amiodarone and alcohol use may indicate 1) anticoagulant use given for the arrhythmia the amiodarone is used for, and 2) coagulopathy caused by chronic alcohol consumption.

While the protective benefits of metformin, GLP-1RA, and bariatric surgery are promising, more therapies for NAFLD/NASH are urgently needed. The mainstay of treatment for NAFLD/NASH is weight loss, and while advances in obesity medicine have made achieving sustainable weight loss more possible,^14^ societal forces including the current Covid-19 pandemic, continue to create an obesogenic environment – over 42% of adults in the US now have obesity.^16^ Research assessing risk from visceral adiposity, such as from imaging studies, is needed.

Our study has several limitations: there may be unmeasured confounders and residual bias. Findings in a sample in the upper Midwest may not be generalizable. Hepatitis B and C were not excluded as they may co-occur with NAFLD/NASH in persons with BMI>=30kg/m^2^, and the prevalence in this region is low: 0.78% of adults with hepatitis C and 0.59% with chronic hepatitis B. Further selection bias may be present in that there are likely many more individuals in this population who have NAFLD/NASH but who did not have 3 elevated ALT readings.^4^

### Conclusions

NAFLD/NASH is a state of chronic inflammation due to visceral adiposity, is a significant risk factor for hospitalization for Covid-19, and appears to account for risk attributed to obesity. Treatments for metabolic disease mitigated risks from NAFLD/NASH. More research is needed to confirm these findings. Patients with elevated BMI should be screened for NAFLD/NASH and informed of the risks associated with visceral adiposity and Covid-19, as well as opportunities for mitigating this risk such as bariatric surgery, GLP-1RA, and metformin.^9,14^

## Data Availability

The data is available in the M Health Fairview Data Cloud

